# MOSAIC: Methylation-Oriented Site Analysis and Information Classifier for Robust Epigenomic Classification of Acute Leukemia in Clinical Cohorts with Variable Tumor Purity

**DOI:** 10.64898/2026.06.16.26355747

**Authors:** Atharva Jaydeep Shah, Donald Green, Lauren Wainman, Jeremiah Karrs, Parth Shah

## Abstract

DNA methylation-based classification offers a rapid diagnostic complement to conventional molecular workflows in acute leukemia. Existing classifiers are trained on array-derived reference cohorts whose construction favors specimens with adequate tumor content, leaving clinically relevant low-purity specimens underrepresented and classifier robustness in this regime uncharacterized. On held-out low-purity specimens, existing classifiers were concordant with expert pathology in only 7 of 10 (MARLIN) and 5 of 10 (ALMA) cases, motivating a classifier built to maintain accuracy at low tumor purity. We developed MOSAIC (Methylation-Oriented Site Analysis and Information Classifier), a neural network classifier built to maintain accuracy across the full range of tumor purities encountered in clinical practice. MOSAIC is a neural network trained on publicly available array-based methylation data augmented with native methylation calls from Oxford Nanopore sequencing. MOSAIC was evaluated on low-purity specimens held out entirely from training. On these held-out low-blast leukemia specimens, all below 25% blasts and including a case at 1.4%, MOSAIC was concordant with expert pathology in every case, recovering the correct subtype where diluted disease signal would otherwise be mistaken for normal or unrelated tissue. Gradient-based saliency analysis showed that the network relies on a partially distinct set of discriminative CpG probes when classifying low-blast specimens. MOSAIC demonstrates that augmenting training with clinically representative clinical specimens yields methylation-based leukemia classification that maintains effectiveness under the variable tumor purity of real clinical cohorts.

## 1 Introduction

Acute leukemia is among the most aggressive hematologic malignancies, demanding rapid and precise molecular classification to guide treatment decisions. The 2022 European LeukemiaNet (ELN) recommendations stipulate that genetic and molecular results should ideally be available within 24 hours to enable risk-adapted therapy selection [1]. In parallel, both the World Health Organization (WHO) Classification of Haematolymphoid Tumours [2] and the International Consensus Classification (ICC) [3] have moved decisively toward genetically defined disease entities, recognizing that morphology alone is insufficient for therapeutic stratification. Of particular relevance to this work, the WHO 2022 classification eliminates the traditional 20% blast requirement for AML with defining genetic abnormalities in many contexts, as patients with lower blast counts and these abnormalities have outcomes resembling those with higher blast counts [2, 4]. A direct corollary is that a specimen with few blasts can nonetheless represent treatment-defining disease, making accurate classification of low-purity specimens a clinical requirement rather than an edge case. Yet current diagnostic workflows remain time-intensive: standard-of-care evaluation requires bone marrow morphology, multiparameter flow cytometry, karyotyping, fluorescence *in situ* hybridization (FISH), and next-generation sequencing panels, a battery that often takes days to weeks and can substantially delay treatment planning [1, 5].

DNA methylation profiling has emerged as a compelling alternative for rapid tumor classification. This approach was pioneered in neuro-oncology, where Capper et al. demonstrated that a machine learning classifier trained on genome-wide methylation profiles could accurately classify central nervous system (CNS) tumors across all entities and age groups, resulting in diagnostic changes in up to 12% of prospective cases [6]. The clinical significance of this work was formalized when the 2021 WHO Classification of CNS Tumors integrated DNA methylation profiling into its diagnostic criteria [7]. Nanopore sequencing further accelerated this paradigm by enabling direct measurement of methylation states from native DNA without bisulfite conversion, supporting same-day genomic and epigenomic diagnosis of brain tumors [8] and intraoperative subclassification from sparse profiles within 40 minutes [9]. These advances established that methylation-based classification, coupled to rapid sequencing, can provide clinically actionable diagnoses on timescales compatible with treatment planning, and the paradigm has since been extended to acute leukemia [10–12].

Existing methylation classifiers for acute leukemia are trained largely on array-derived reference cohorts, whose construction favors specimens with adequate tumor content for hybridization. This leaves low-purity specimens underrepresented in the training distribution and classifier robustness below this implicit purity floor uncharacterized. Low purity degrades classification through a concrete mechanism: bulk samples contain mixed neoplastic, immune, and stromal populations, allowing admixed normal methylation signals to alter beta values and confound classification [13–15]. In acute leukemia, blast percentage spans from over 90% to under 5% [16]; at the low end, normal hematopoietic methylation dominates and dilutes the disease-specific signal on which the classifier depends. A classifier whose training distribution excludes this regime is therefore vulnerable to dataset shift [17, 18] encountered when it is deployed on unselected clinical specimens, precisely the low-purity cases that recent classification criteria make diagnostically essential.

We developed MOSAIC (Methylation-Oriented Site Analysis and Information Classifier), a neural network classifier built to maintain accuracy across the full range of tumor purities encountered in clinical practice. MOSAIC uses a 512-node first hidden layer and augments training with class-balanced clinical specimens from our institution. Wider networks improve generalization when effective task complexity rises, provided sufficient training data to avoid overfitting [19, 20]. We present the design and evaluation of MOSAIC, a classifier trained to be robust to the tumor purity variation that characterizes unselected clinical cohorts, and assess its performance against existing classifiers in the low-purity regime.

## 2 Methods

### 2.1 Clinical cohort and data preprocessing

Sixty-three samples were collected from patients evaluated at Dartmouth-Hitchcock Medical Center (DHMC) under institutional review board-approved protocols, comprising 62 AML bone marrow aspirates and one normal bone marrow control. All samples were sequenced on the Oxford Nanopore Technologies PromethION 2 Solo or PromethION 24 platform, and methylation calls were extracted using Modkit (v 0.3.3), producing a methylbed file containing per-CpG beta values. Clinical diagnoses were established through standard-of-care workup (flow cytometry, cytogenetics, FISH, gene panel sequencing) and expert pathology review. Blast percentages were recorded from clinical flow cytometry and/or morphologic assessment. Disease sub-types were defined according to the methylation classification scheme of as previously described [10].

Of the 63 samples, 53 had expert pathology-confirmed methylation class annotations spanning 18 distinct subtypes; the remaining 10 lacked definitive annotations. Three of the annotated cases were additionally analyzed at multiple time points for real-time trajectory analysis (Sec. 3.3). The cohort exhibited substantial heterogeneity in tumor purity, with blast counts among annotated samples ranging from 0% (normal bone marrow control) to 95% (median 52.5%).

To evaluate generalization to low-purity material, all annotated specimens with blast counts below 25% were withheld from training entirely and reserved as a held-out evaluation set, comprising 12 unique specimens. The remaining 40 annotated specimens were used for training augmentation. Of the 12 held-out specimens, one carried an ambiguous dual-class annotation for which no single ground-truth label could be assigned and was excluded from strict scoring, leaving 11 strict-evaluable specimens: 10 low-blast leukemia cases (including three with blasts *<*5%, the lowest at 1.4%) and one normal bone marrow control (0% blasts). Because every held-out specimen was excluded from training entirely, performance on this set reflects out-of-sample generalization to the low-purity regime (Sec. 3.2).

Methylbed files were parsed to extract per-CpG beta values and CpG identifiers. Data were pivoted to a sample-by-CpG matrix and aligned to the 357,340 reference CpG probe positions to allow backward compatibility with historical methylation data derived from arrays. Missing features were imputed with *β* = 0.5 (the binarization boundary). Beta values were binarized to +1 (≥ 0.5) and − 1 (*<* 0.5). The reference dataset (2,356 samples, 42 classes) was augmented with the 40 high-purity annotated DHMC specimens and peripheral blood control subclasses were merged. For each class, 50 samples were drawn with replacement, and 10% of CpG sites were randomly flipped to simulate noise.

### 2.2 Model architecture and training

MOSAIC takes the 357,340 reference CpG positions as input and passes them through two hidden layers, the first with 512 sigmoid units and the second with 128 ReLU units, before a 42-way softmax output. During training, 99% input dropout is applied at each epoch. In total the network has 183,029,674 parameters (Table 1).

**Table 1.** MOSAIC architecture.

| Component | Configuration |
| --- | --- |
| Input layer | 357,340 CpG positions |
| Hidden layer 1 | 512 nodes, sigmoid |
| Hidden layer 2 | 128 nodes, ReLU |
| Output layer | 42 nodes, softmax |
| Input dropout | 0.99 (training only) |
| Batch size | 128 |
| Total parameters | 183,029,674 |

The width of the first hidden layer was selected by a sweep over candidate sizes (128, 256, 384, 512, 640, and 768 nodes), holding all other hyperparameters fixed. Validation concordance on held-out low-purity specimens was highest at 512 nodes; narrower layers (128, 256, 384) gave lower accuracy on low-blast specimens, and wider layers (640, 768) gave no further improvement. The 512-node configuration was therefore selected.

The model was trained for 3,000 epochs using the Adam optimizer (learning rate 10^−5^) [21], categorical cross-entropy loss, and a batch size of 128 (Fig. 2). Random seeds were fixed for reproducibility. The reference cohort beta values were binarized as described in Sec. 2.1. Predictions were generated from softmax probabilities; the predicted class is the highest-probability class and the confidence score is that maximum probability. Predictions with confidence ≥ 0.8 were considered high-confidence [10].

### 2.3 Interpretability analysis

To identify the CpG probes most influential for each classification decision, we computed input-level gradient saliency maps following the method of Simonyan et al. [22]. For each sample, the gradient of the predicted class logit with respect to the 357,340 input features was computed by backpropagation through the trained network. For a sample **x** with predicted class *c*, the saliency at CpG position *i* is |∂*z*_*c*_*/*∂*x*_*i*_|, where *z*_*c*_ is the pre-softmax logit for class *c*. The chain rule through the network layers gives:

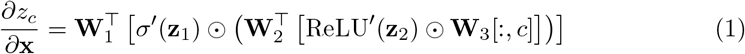

where **W**_1_, **W**_2_, **W**_3_ are the weight matrices, *σ*^*′*^ and ReLU^*′*^ are the activation derivatives, and ⊙ denotes element-wise multiplication. Dropout is inactive at inference, so the saliency map is a deterministic function of the input. Class-level saliency profiles were computed by averaging per-sample saliency maps across all samples predicted to belong to each class. Differential saliency between high-blast (≥ 25%) and low-blast (*<*25%) specimens was computed to identify CpG probes whose importance shifts with tumor purity.

To characterize how the wider architecture redistributes feature utilization relative to the 256-node network as previously described in [10], we compared their first-layer weight matrices (357,340 *×* 256 and 357,340 *×* 512, respectively). For each model, per-CpG importance was computed as the mean absolute first-layer weight across all hidden units, normalized to [0, 1]. Because the two networks differ in hidden-layer width and initialization, this comparison characterizes relative shifts in per-CpG emphasis rather than unit-level correspondence.

## 3 Results

### 3.1 Cohort characterization and training dynamics

The annotated DHMC cohort (*n* = 53) encompassed 18 methylation classes, most frequently Chromatin/Spliceosome-enriched (*n* = 14), TP53/Aneuploidy-enriched (*n* = 12), and HOX-activated subgroups (*n* = 12) based on the molecular findings. The blast-count distribution across the full annotated cohort (Fig. 1) shows enrichment of low-blast specimens relative to array-derived research cohorts; 13 specimens (24.5%) had blast counts below 25% and constituted the held-out evaluation set (Sec. 2.1). Training on the augmented dataset converged within ∼ 500 epochs (Fig. 2): categorical cross-entropy loss decreased from ∼ 3.9 to 0.029, and training precision and recall reached 0.998 by epochs 300 and 500, respectively, remaining stable through 3,000 epochs. Figure 2 shows training convergence; out-of-sample performance is reported in Sec. 3.2.

**Fig. 1.**
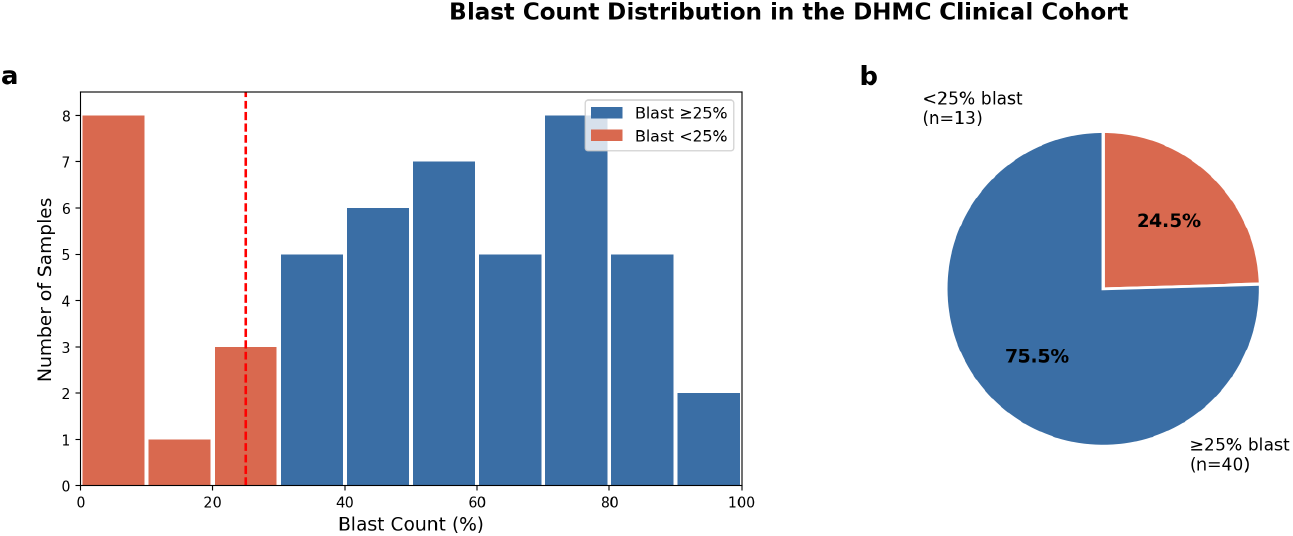
Blast count distribution in the full annotated DHMC cohort. (a) Histogram of blast percentages across the 53 annotated samples. Red bars: blast *<*25%; blue bars: blast ≥25%; dashed line: 25% threshold. (b) Proportion of samples below versus at or above ≥ 25% blasts. The low-blast group comprises 12 unique specimens; the normal bone marrow control was recorded as two sequencing files, giving 13 records.

**Fig. 2.**
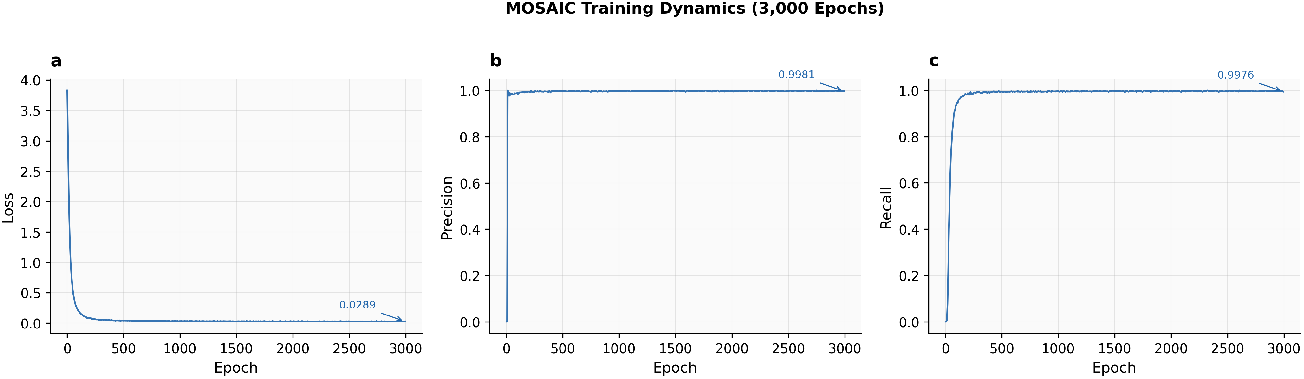
MOSAIC training dynamics (3,000 epochs). (a) Categorical cross-entropy loss. (b) Training precision. (c) Training recall. Final values annotated.

### 3.2 Held-out low-purity performance and classifier comparison

The primary evaluation was performed on the low-purity specimens withheld entirely from training (Sec. 2.1). Of the 12 unique held-out specimens, 10 were strict-evaluable leukemia cases (all *<*25% blasts); one of the specimen was normal bone marrow control and one specimen (Sample 49, 9.7% blasts) had an ambiguous dual-class annotation which was excluded from the leukemia concordance comparison.

On the 10 strict-evaluable held-out leukemia specimens, MOSAIC was concordant with expert pathology in every case (10/10), with a mean confidence of 0.995 (Table 3, Fig. 3). Concordance was maintained at the lowest tumor purities, including a specimen with 1.4% blasts classified correctly above the high-confidence threshold. The held-out normal bone marrow control (Sample 28, 0% blasts), was assigned to a control class rather than a malignant class, indicating that the network does not force a leukemic prediction on normal marrow.

**Fig. 3.**
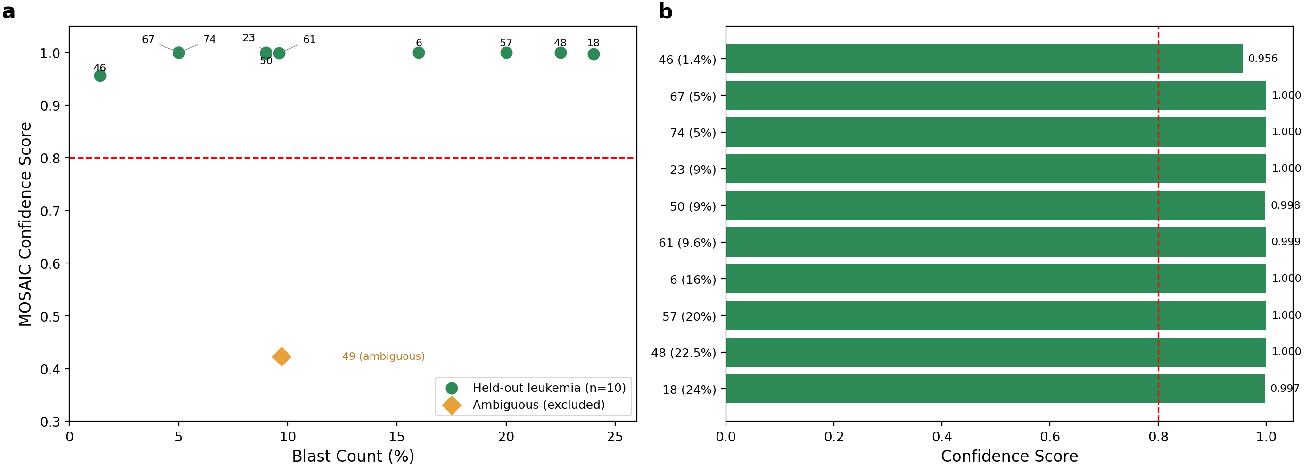
Performance on held-out low-blast specimens (*<*25% blasts). (a) Blast count vs. confidence score. (b) Per-sample confidence bars ordered by blast count.

On the same out-of-sample specimens, the pretrained MARLIN checkpoint [10] was concordant in 7 of 10 (7/10) and ALMA [11] in 5 of 10 (5/10) (Table 2). Because no held-out specimen contributed to MOSAIC’s training, all three classifiers were evaluated out-of-sample on this set, making the comparison directly interpretable. The discordant MARLIN calls were concentrated in the lowest-purity specimens: two 5%-blast specimens were assigned to a T-cell class and a HOX class, respectively, and a 24%-blast specimen was assigned to a peripheral-blood control class. In each case the diluted disease signal led the reference-trained classifier toward a normal-cell or unrelated prediction, whereas MOSAIC recovered the expected class. All classifiers received identical binarized inputs, consistent with low tumor purity rather than sequencing platform as the operative factor.

**Table 2.** Concordance with expert pathology on the held-out low-purity leukemia specimens (*n* = 10, all *<*25% blasts). All three classifiers were evaluated out-of-sample on this set.

| Classifier | Concordant / total |
| --- | --- |
| MOSAIC | 10/10 |
| MARLIN | 7/10 |
| ALMA | 5/10 |

**Table 3.** MOSAIC classification of held-out low-blast specimens (*<*25% blasts). The normal marrow control and one ambiguous specimen are shown separately and excluded from the strict leukemia concordance (*n* = 10).

| SampleID | Blast (%) | True Class | MOSAIC Prediction | Confidence |
| --- | --- | --- | --- | --- |
| 46 | $<5$ | Chromatin/Spliceosome | Chromatin/Spliceosome | 0.956 |
| 67 | $<5$ | Chromatin/Spliceosome | Chromatin/Spliceosome | 1.000 |
| 74 | $<5$ | Chromatin/Spliceosome | Chromatin/Spliceosome | 1.000 |
| 23 | 9 | Chromatin/Spliceosome | Chromatin/Spliceosome | 1.000 |
| 50 | 9 | Chromatin/Spliceosome | Chromatin/Spliceosome | 0.998 |
| 61 | 9.6 | Chromatin/Spliceosome | Chromatin/Spliceosome | 0.999 |
| 6 | 16 | Chromatin/Spliceosome | Chromatin/Spliceosome | 1.000 |
| 57 | 20 | CEBPA | CEBPA | 1.000 |
| 48 | 22.5 | HOX Grp 5 | HOX Grp 5 | 1.000 |
| 18 | 24 | IDH1/2 | IDH1/2 | 0.997 |
| *Excluded from strict leukemia concordance:* |  |  |  |  |
| 28 | 0 | BM control | BM control | 1.000 |
| 49 | 9.7 | Ambiguous (dual-class) | — | — |

To visualize the relationship between DHMC specimens and the reference cohort in methylation feature space, we computed a joint t-SNE embedding of all 2,419 samples (2,356 reference and 63 DHMC; Fig. 4). Each DHMC specimen is colocalized with the reference cluster corresponding to its assigned subtype. MDS-associated and TP53/Aneuploidy-related entities occupied closely neighboring regions of the embedding, consistent with the recognized biological overlap among these myelodysplasia-related classes.

**Fig. 4.**
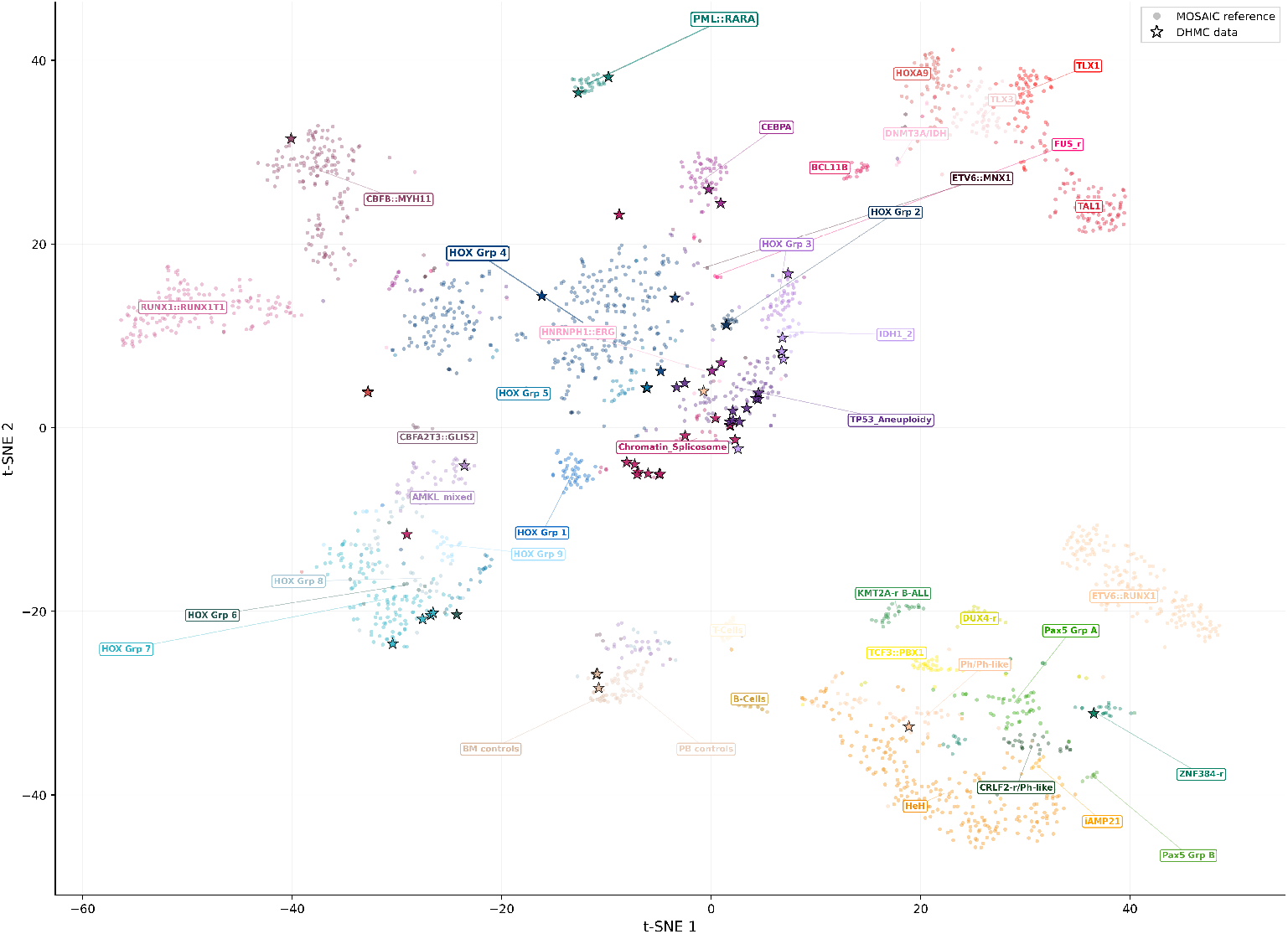
t-SNE visualization of the MOSAIC methylation landscape. Unsupervised t-SNE embedding of 2,419 samples (2,356 reference and 63 DHMC), computed from binarized methylation features after PCA pre-reduction to 50 components. Small colored points indicate reference samples; starred points indicate DHMC clinical samples. Class names are placed at reference-cluster centroids. DHMC samples co-localize with reference samples of their assigned subtype.

### 3.3 Real-time classification trajectory

We assessed real-time classification on time-point data from three prospective cases (Fig. 5). In the first case, an initial HOX-group prediction transitioned to PML::RARA by approximately 2 hours and reached confidence 1.00 by 4 hours, remaining stable thereafter. In the second case, the classifier reached the correct KMT2A-r B-ALL prediction at confidence 1.00 from the first time point and maintained it across all time points over 18 hours. In the third case, IDH1/2 emerged as the leading prediction by approximately 2 hours and remained the top call thereafter, though confidence oscillated against HOX Grp 3 and stabilized at 0.87 only by 18 hours, the expected behavior when sequencing depth remains marginal for a difficult-to-classify specimen. Across these cases, MOSAIC converged to the correct high-confidence prediction within 2– 4 hours for specimens with adequate coverage and reported lower confidence where coverage remained insufficient.

**Fig. 5.**
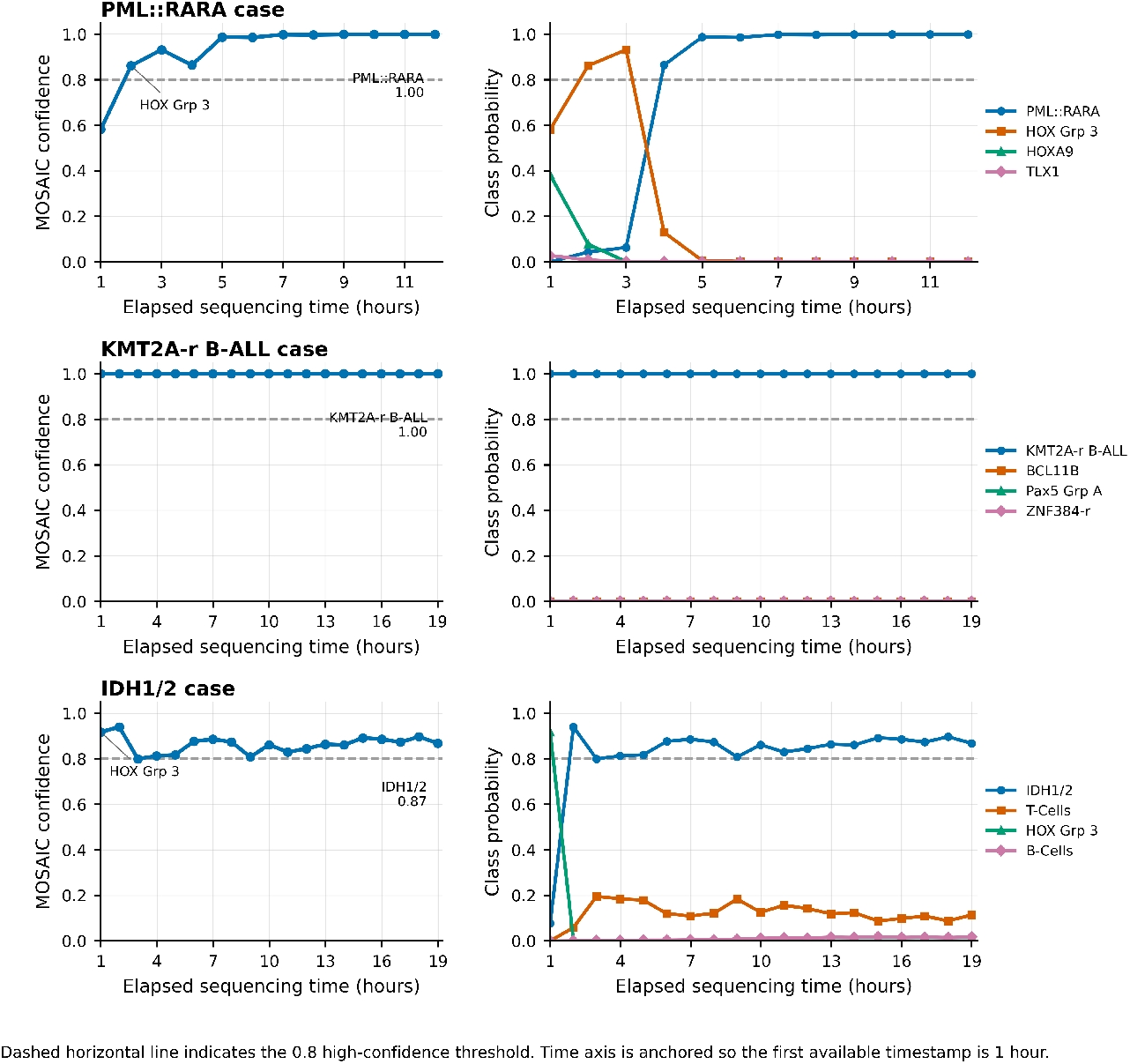
Real-time classification trajectories for three prospective cases. Each row is one sequencing run. Left panels show MOSAIC confidence over elapsed sequencing time, with the leading predicted class annotated; right panels show class-probability trajectories for the top competing classes. The dashed line indicates the 0.8 high-confidence threshold. Top: convergence to PML::RARA within approximately 2–4 hours. Middle: immediate stable classification as KMT2A-r B-ALL. Bottom: oscillation between IDH1/2 and HOX Grp 3, reflecting marginal sequencing depth.

### 3.4 Class-discriminative CpG probes

Gradient saliency analysis identified distinct sets of CpG probes driving classification for each leukemia subtype (Figs. 6 and A1). Each subtype was characterized by a distinct saliency profile, with the top discriminative probes per class showing substantially elevated importance relative to the genome-wide background. For example, the Chromatin/Spliceosome class was most strongly influenced by probes cg18176482, cg14720960, and cg05011848; TP53/Aneuploidy by cg15230030, cg24731066, and cg25770481; and IDH1/2 by cg20539142 and cg07660102. The cross-class saliency heatmap (Fig. 6) indicated that discriminative CpG sets are largely non-overlapping between subtypes, consistent with the biological distinctiveness of these leukemia entities. Here, CpG sites are referred to by their Illumina Infinium methylation BeadChip probe identifiers (the cg-prefixed IDs from the HumanMethylation450 and MethylationEPIC arrays), each denoting a single genomic CpG locus.

**Fig. 6.**
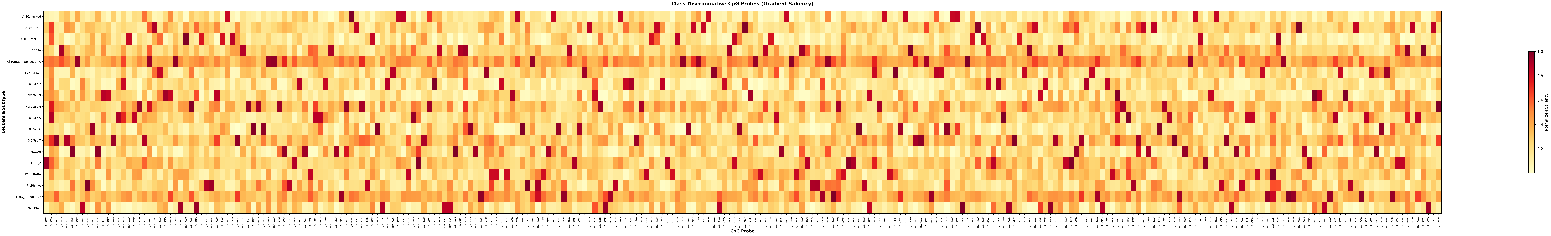
Class-discriminative CpG saliency heatmap. Gradient saliency scores for the top 15 discriminative CpG probes per leukemia subtype. Rows represent subtypes; columns represent CpG probes; color intensity reflects normalized saliency.

Across all samples and classes, genome-wide saliency analysis ranked all 357,340 CpG probes by mean importance; we highlight the 100 highest-scoring probes (Fig. 7). The distribution of saliency scores was highly skewed, with the majority of probes contributed minimally to classification, while a small subset carried disproportionate discriminative weight, indicating that the network relies on a compact feature set despite the high dimensionality of the input space.

**Fig. 7.**
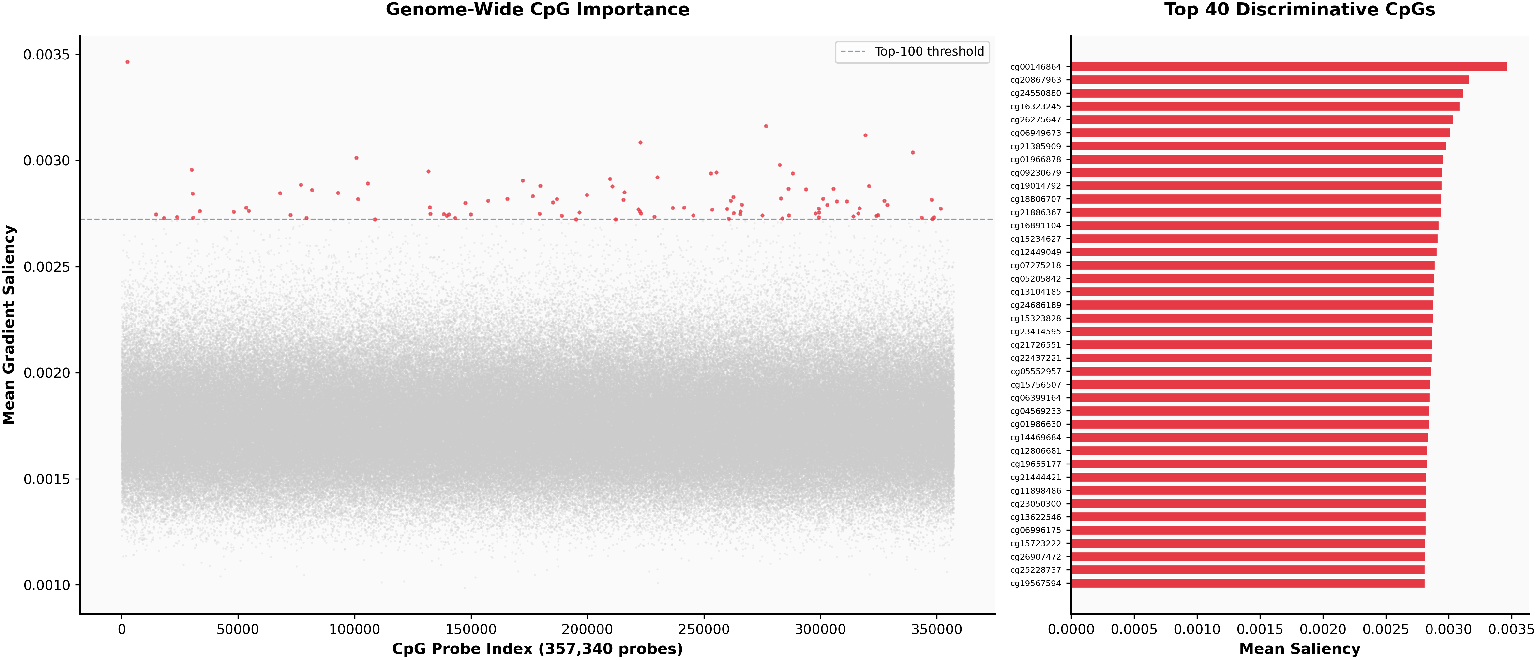
Genome-wide CpG importance. (a) Manhattan-style plot of mean gradient saliency across all 357,340 CpG probes. Red: probes exceeding the top-100 importance threshold. (b) Top 40 discriminative CpG probes ranked by mean saliency score.

### 3.5 Differential CpG importance by tumor purity

Comparing saliency between high-blast (≥ 25%) and low-blast (*<*25%) specimens revealed a shift in CpG utilization with tumor purity (Fig. 8). Thirty CpG probes showed substantially increased saliency in low-blast specimens relative to high-blast specimens, led by cg05139728, cg10781663, and cg24793022, with all top-30 differentially important probes enriched in the low-blast direction. This indicates that the network draws on a partially distinct feature set when tumor-derived signal is diluted by normal hematopoietic methylation, consistent with the model maintaining classification by attending to probes that retain discriminative information at low tumor purity.

**Fig. 8.**
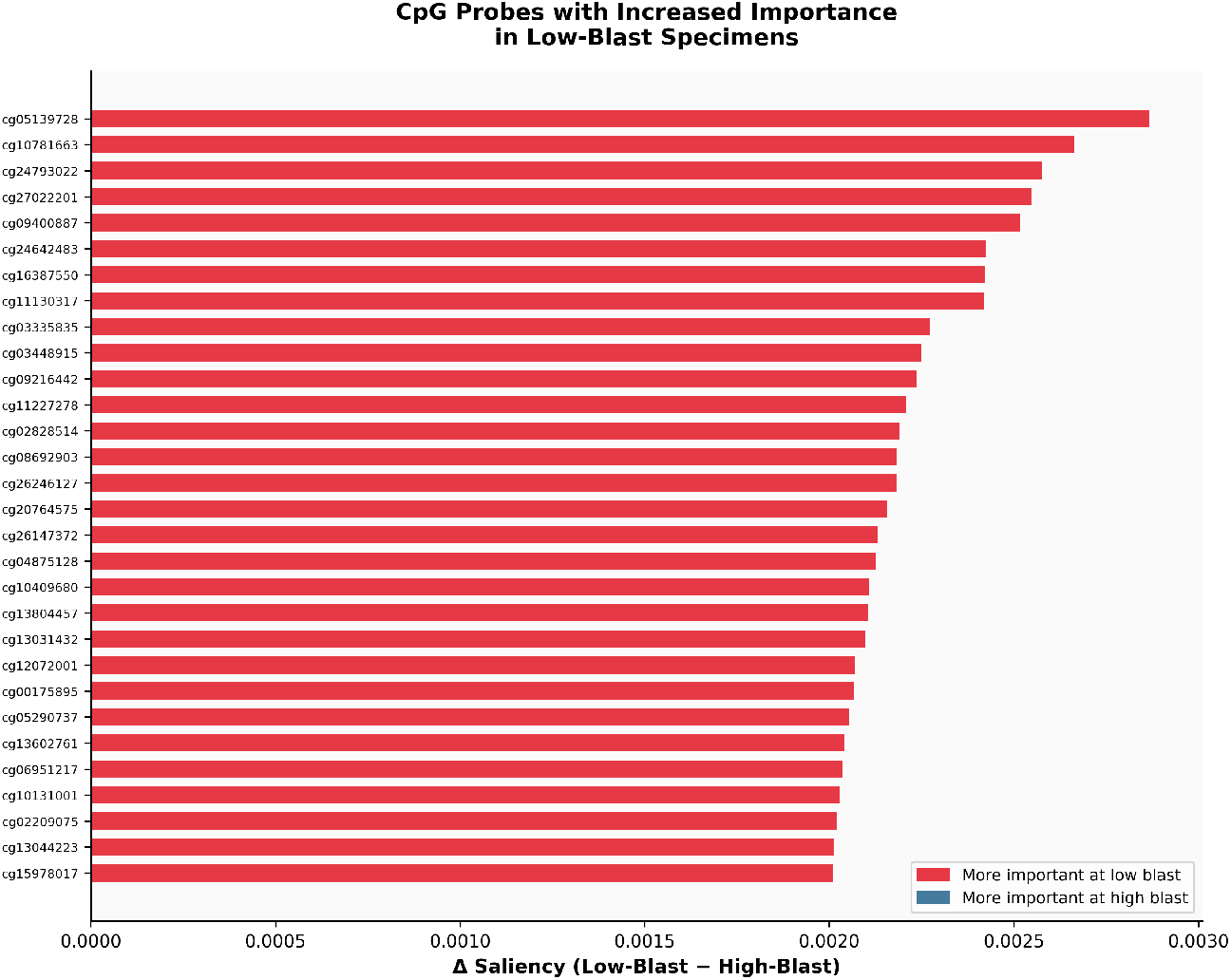
Differential CpG importance by tumor purity. CpG probes ranked by the difference in gradient saliency between low-blast (*<*25%) and high-blast (≥ 25%) specimens. All top-30 differentially important probes are enriched in the low-blast direction.

**Fig. 9.**
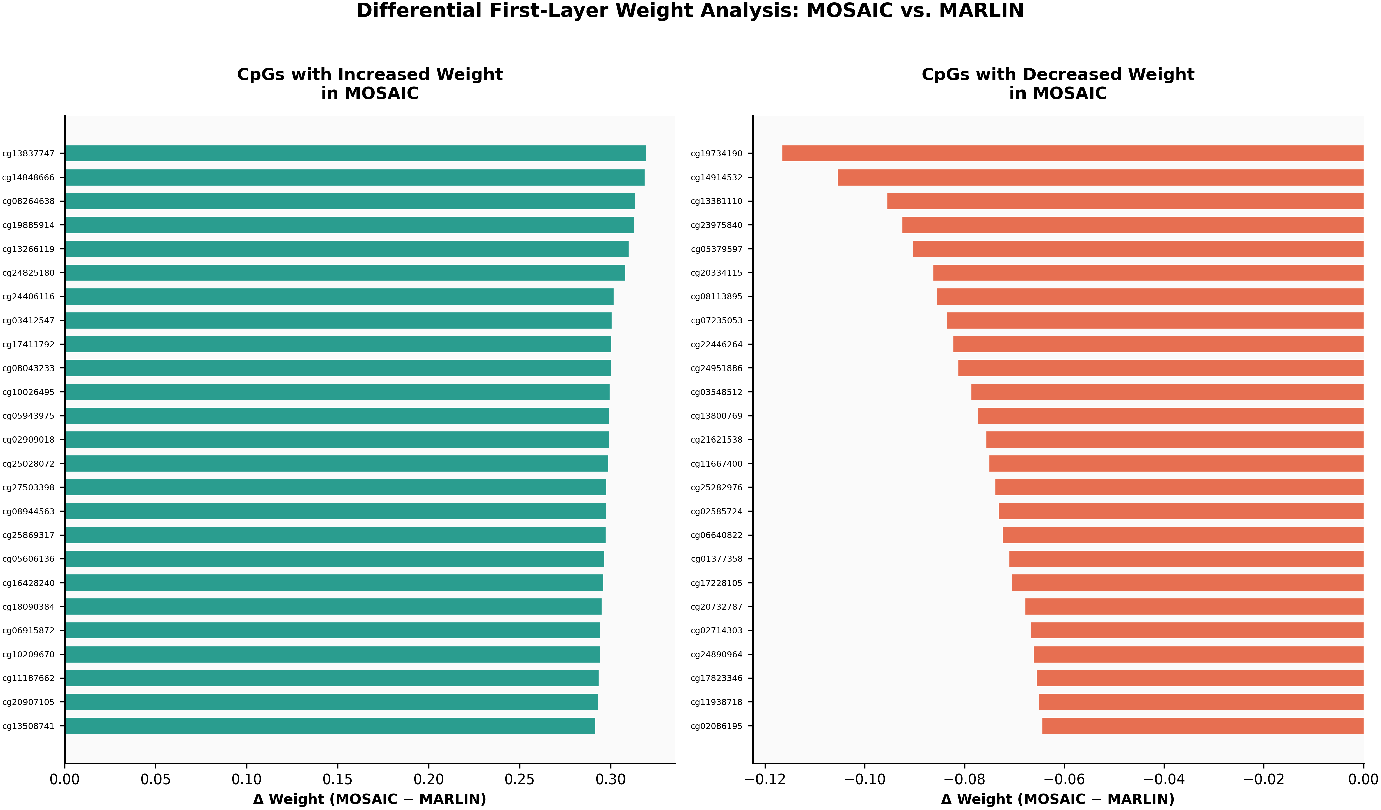
Differential first-layer weight analysis: MOSAIC vs. the 256-node base architecture (MAR-LIN). (a) CpG probes with the largest increase in normalized first-layer weight magnitude in MOSAIC. (b) CpG probes with the largest decrease.

### 3.6 Differential first-layer weight utilization

We compared first-layer weight magnitudes between MOSAIC and the 256-node architecture described in [10] (9). Twenty-five CpG probes showed the largest increases in normalized weight magnitude in MOSAIC, and a distinct set of 25 probes showed the largest decreases 3.4. Because the two networks differ in hidden-layer width and initialization, this comparison characterizes relative shifts in per-CpG emphasis rather than unit-level correspondence; the bidirectional redistribution indicates that the wider network does not simply scale the base feature weighting but emphasizes a partially distinct set of probes.

## 4 Discussion

The central challenge addressed here is a dataset shift that broadly affects clinical AI deployment: models trained on curated research data often degrade when applied to clinical specimens that differ systematically from the training distribution [17, 18, 23]. This shift has a concrete and measurable cause here. Array-derived reference cohorts require sufficient tumor content for hybridization, so the low-purity regime is structurally absent from training, and a classifier never exposed to diluted specimens has no basis for handling them. Augmenting training with clinically representative specimens narrows that gap: on low-purity specimens withheld entirely from training, MOSAIC classified every low-blast leukemia case concordantly with expert pathology, including one at 1.4% blasts, recovering subtypes that reference-trained classifiers assigned to normal-cell or unrelated classes. This mirrors the broader finding that site-specific retraining improves cross-institutional performance [24, 25], and identifies tumor purity as the specific axis along which that retraining matters for leukemia methylation classifiers.

Saliency analysis indicates how the model sustains accuracy as purity falls. Each subtype mapped to a distinct, largely non-overlapping set of discriminative CpG probes, and at low blast counts the network shifted toward a partially distinct probe set, drawing on features that remain informative once tumor signal is diluted by normal hematopoietic methylation. We frame this as a characterization of the trained model rather than a controlled test of mechanism: isolating the architectural or training factors responsible would require targeted ablations, which we did not perform.

The class-specific discriminative probes also have potential biological relevance. Fewer than 0.03% of the 357,340 probes carried the majority of discriminative weight, suggesting the model concentrates on a compact feature set. Mapping these probes to annotated genomic features and cross-referencing known methylation alterations in AML subtypes is a natural direction for biological validation and for identifying candidate subtype-specific epigenetic markers.

From a clinical standpoint, MOSAIC addresses a gap that current classification criteria make increasingly relevant. The 2022 WHO and ICC classifications recognize many genetically defined AML entities at blast counts well below the traditional 20% threshold [2–4], so a classifier that degrades at low blast counts is at odds with how disease is now defined. Maintaining accuracy across the full blast range, including specimens as low as 1.4% blasts, positions methylation-based classification for settings where tumor purity cannot be controlled or pre-selected.

Several limitations should be acknowledged. Evaluation was performed at a single site, and the held-out low-purity set is small (10 strict-evaluable leukemia specimens); multi-center validation across diverse populations is an essential next step for establishing generalizability and supporting clinical deployment. A natural route to that validation is a federated approach, in which a single classifier is trained across multiple institutions without centralizing patient data, broadening the represented purity and subtype distribution while respecting site-level data governance. The softmax probabilities used as confidence scores are not calibrated posterior probabilities. Finally, the saliency and weight analyses are descriptive characterizations of the trained model rather than controlled tests of mechanism, and the specific architectural configuration selected here may not transfer directly to cohorts with different subtype distributions or sequencing depths. Future work will include evaluation against non-AML hematologic malignancies to assess broader generalizability.

## 5 Conclusion

Methylation-based classifiers hold considerable promise for rapid leukemia diagnostics, but their clinical utility depends on robustness to the variable specimen quality encountered outside curated research settings. MOSAIC extends classification reliably into the low-purity regime: on specimens withheld entirely from training, it classified every low-blast leukemia case concordantly with expert pathology, including a specimen with 1.4% blasts that would challenge conventional diagnostic workflows. Gradient-based interpretability analysis further shows that subtype predictions are traceable to a compact, largely non-overlapping set of discriminative CpG probes, and that the model’s feature usage shifts as tumor purity decreases, offering a basis for future biological investigation into subtype-specific epigenetic markers. As classification systems continue to move toward genetically defined disease entities with relaxed blast thresholds, diagnostic tools must keep pace. MOSAIC offers a step in that direction, and its architecture provides a foundation for extension to additional leukemia lineages, multi-center validation, and integration with complementary molecular assays.

## Data Availability

All data produced in the present study are available upon reasonable request to the authors

## Acknowledgements

The authors thank the clinical and laboratory teams at Dartmouth-Hitchcock Medical Center for sample collection and pathology review.

## Declarations

### Ethics approval

Samples were collected under institutional review board-approved protocols at Dartmouth-Hitchcock Medical Center.

### Consent for publication

Not applicable.

### Data availability

DHMC clinical data are available upon reasonable request subject to institutional data sharing agreements. The MARLIN reference cohort is available from Steinicke, Benfatto et al. [10].

### Code availability

Code is not publicly available due to the presence of patient-identifiable information in the processing pipeline. Requests for access may be directed to the corresponding author.

### Author contribution

A.J.S. conceived and designed the study, developed the MOSAIC architecture, performed computational analyses, generated figures, and wrote the manuscript. D.G. provided clinical samples and pathology expertise. L.W. contributed to laboratory workflow coordination, sequencing data generation, and manuscript revision. J.K. contributed to sample coordination, clinical data curation, pathology expertise and manuscript revision. P.S. supervised the study, contributed to data interpretation, and revised the manuscript. All authors read and approved the final manuscript.

## Appendix A Supplementary figures

**Fig. A1.**
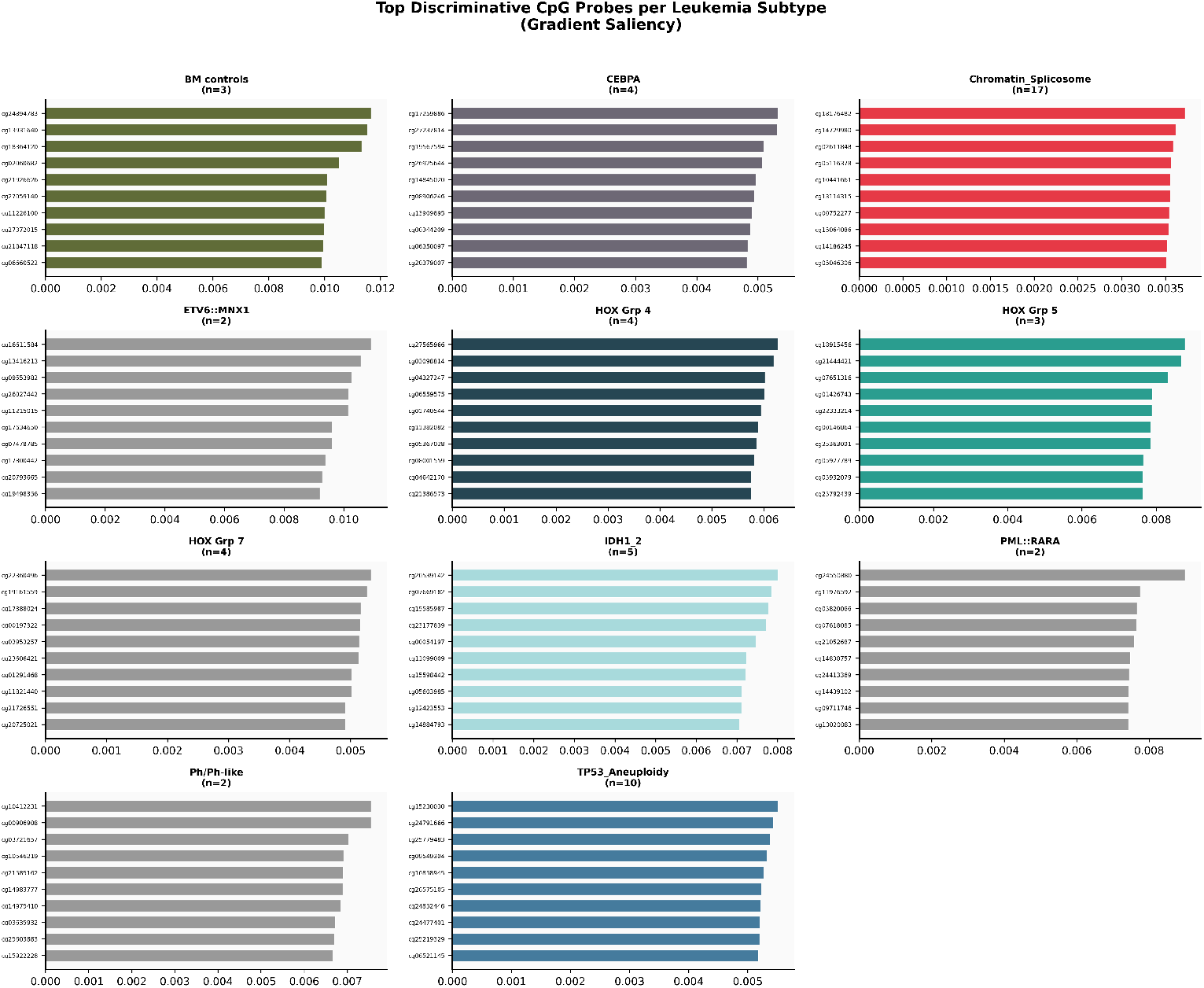
Top discriminative CpG probes per leukemia subtype. Grid of bar charts showing the 10 most important CpG probes (by gradient saliency) for each subtype with ≥ 2 samples. Subtypes are color-coded; sample counts annotated.

